# Incremental Prognostic Value of Regional Myocardial Flow Reserve Beyond Global Flow and Perfusion: REFINE PET

**DOI:** 10.64898/2026.10.29.26366834

**Authors:** Hidesato Fujito, Donghee Han, Alan Rozanski, Gloria J. Hong, Giselle Ramirez, Joanna X. Liang, Mark Lemley, Aakash Shanbhag, Robert J.H. Miller, Paul B. Kavanagh, Heidi Gransar, Leandro Slipczuk, Mark I. Travin, Erick Alexanderson, Isabel Carvajal-Juarez, Rene R.S. Packard, Mouaz Al-Mallah, Andrew J. Einstein, Attila Feher, Wanda Acampa, Stacey Knight, Viet T. Le, Steve Mason, Panithaya Chareonthaitawee, Ronny R. Buechel, Thomas L. Rosamond, Robert A. deKemp, Damini Dey, Marcelo F. Di Carli, Daniel S. Berman, Piotr J. Slomka

**Author notes:** Donghee Han, MD and Piotr J. Slomka, PhD contributed equally as co-corresponding authors. **Corresponding author:** Donghee Han, M.D. Department of Cardiology, Smidt Heart Institute, Cedars-Sinai Medical Center 8700 Beverly Blvd. Los Angeles, CA 90048, Piotr J. Slomka, PhD Department of Medicine, Cedars-Sinai Medical Center 8700 Beverly Blvd. Los Angeles, CA 90048.

## Abstract

**Background:** The incremental prognostic value of regional myocardial flow reserve (MFR) assessed by positron emission tomography (PET) myocardial perfusion imaging (MPI) beyond global MFR remains unclear. We sought to evaluate the prognostic value of regional MFR in the international multicenter REFINE PET registry.

**Methods:** This retrospective multicenter cohort study included 35,391 patients from 13 sites in the international REFINE PET registry who underwent rubidium-82 (^82^Rb) and ^13^N-ammonia (^13^NH_3_) PET MPI. Abnormal MFR was defined as <2.0. Patients were classified into three groups: MFR-Group 1 (normal global and regional MFR; n=19,744), MFR-Group 2 (normal global MFR but abnormal regional MFR; n=3,184), and MFR-Group 3 (abnormal global and regional MFR; n=12,463). Major adverse cardiovascular events (MACE) included all-cause mortality, non-fatal myocardial infarction, late revascularization (>90 days), and hospital admission for unstable angina.

**Results:** During a mean follow-up of 4.0 ± 2.9 years, 9,903 patients (28.0%) experienced MACE. Annualized MACE rates increased stepwise across MFR-Groups 1-3 (4.0%/year, 7.5%/year, and 11.8%/year, respectively; *p*<0.001). Abnormal regional MFR independently predicted MACE after adjustment for clinical risk factors, global MFR, and stress total perfusion deficit (HR 1.34, 95% CI 1.25–1.43). Adding regional MFR to a model including global MFR significantly improved model fit (χ² 4,784 to 4,871; *p*<0.001), discrimination (AUC 0.710 to 0.729; *p*<0.001), and reclassification (NRI 41.9%). Among patients with normal perfusion and preserved global MFR, abnormal regional MFR identified higher risk (5.0%/year vs. 2.8%/year; *p*<0.001).

**Conclusions:** Regional MFR provided incremental prognostic value beyond conventional PET MPI findings, including global MFR, and may refine risk stratification, particularly when global MFR is preserved.

**Clinical perspectives:** Global myocardial flow reserve (MFR) is widely used for risk stratification in patients undergoing positron emission tomography (PET) myocardial perfusion imaging (MPI). However, regional abnormalities in MFR may be overlooked when global MFR is preserved. In this international multicenter study of 35,391 patients, abnormal regional MFR was independently associated with major adverse cardiovascular events and provided incremental prognostic value beyond conventional clinical and PET-MPI findings, including global MFR and myocardial perfusion abnormalities. Importantly, even among patients with normal perfusion and preserved global MFR, those with abnormal regional MFR had a substantially higher annualized event rate than those with normal regional MFR (5.0% versus 2.8%). These findings highlight the importance of assessing regional MFR in addition to global MFR during routine PET-MPI interpretation.

**Graphic Abstract:** 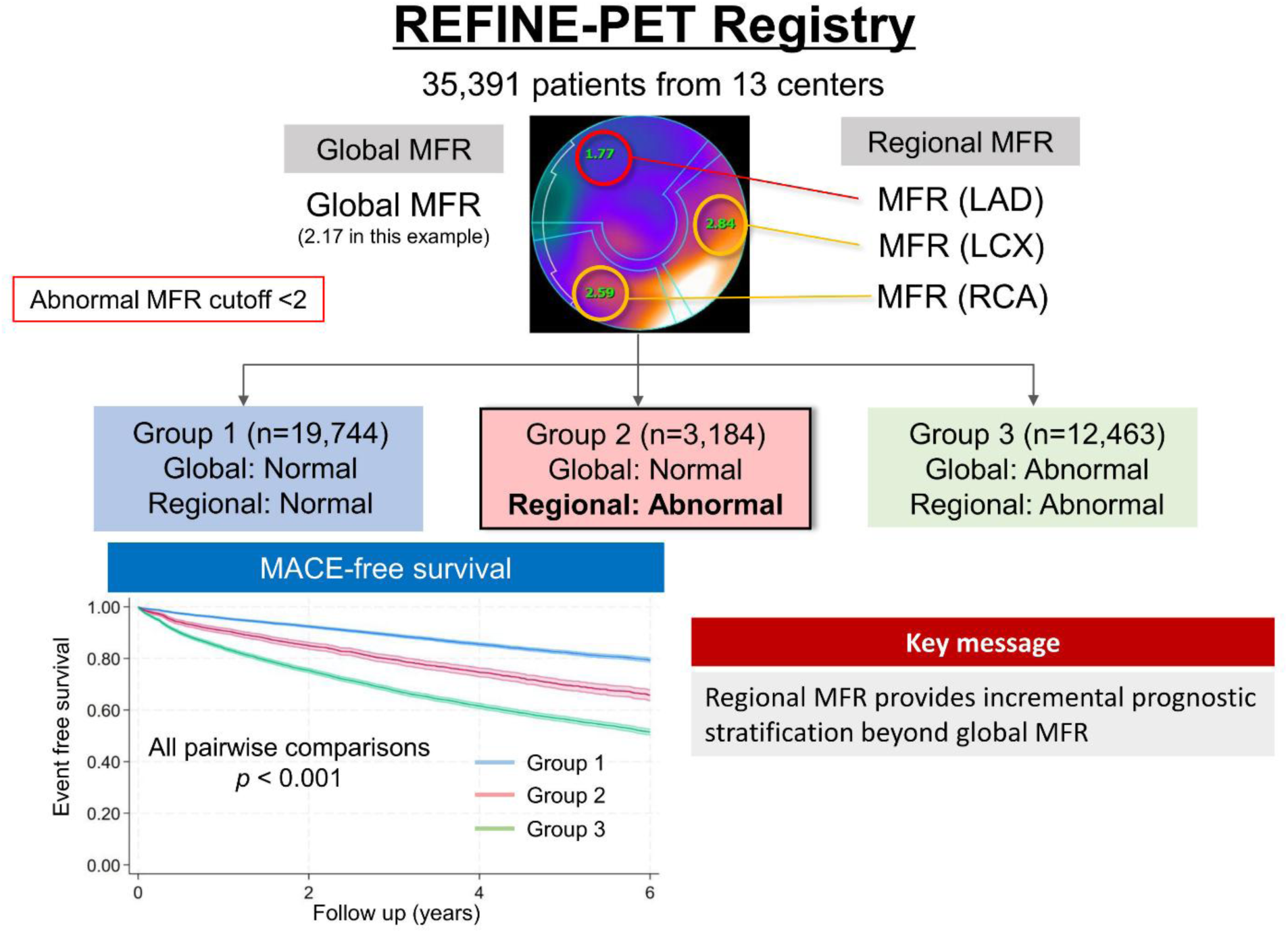

## Introduction

Positron emission tomography (PET) myocardial perfusion imaging (MPI) is a cornerstone test for diagnosis of coronary artery disease (CAD) ^1^. Beyond detecting myocardial perfusion defects, PET-MPI enables the quantification of myocardial flow reserve (MFR), defined as the ratio of myocardial blood flow (MBF) during maximal vasodilation to resting MBF ^2^. This assessment provides a comprehensive physiological evaluation of flow-limiting disease across the coronary vasculature, establishing MFR as a valuable diagnostic and prognostic marker ^2–5^. Global MFR, which averages the blood flow reserve across the entire myocardium, is the standard imaging measurement in PET-MPI for assessing multi-vessel diseases or microcirculatory dysfunction ^1,5,6^. However, focal coronary artery lesions or microvascular dysfunction may only result in subtle regional difference in MFR without alteration of global MFR^7^. PET-MPI also allows assessment of MBF and MFR at the segmental or coronary territory level. Despite this capability, the independent and additive prognostic value of regional MFR remains largely unexplored, particularly among patients with otherwise reassuring PETMPI findings, including preserved global MFR and minimal perfusion abnormalities. Regional MFR may help identify higher-risk patients within this population. Accordingly, the present study aimed to comprehensively evaluate the prognostic value of regional MFR beyond conventional PET-MPI findings using rubidium-82 (^82^Rb) and ^13^N-ammonia (^13^NH_3_) PET MPI from the REgistry of Flow and Perfusion Imaging for Artificial INtelligEnce with PET (REFINE PET) registry.

## Methods

The deidentified data and analysis code underlying this study may be made available upon reasonable request, subject to applicable data-sharing agreements and institutional review board approvals.

### Study population

This retrospective multicenter study included 35,391 consecutive patients with suspected or established CAD who underwent PET MPI at 13 REFINE PET registry sites between 2007 and 2024 ^8^. All data were anonymized prior to transfer using dedicated deidentification software fully compliant with the Health Insurance Portability and Accountability Act (HIPAA). The study protocol was approved by the institutional review boards of all participating sites, and overall study oversight was provided by the Cedars Sinai Medical Center institutional review board.

### Clinical data

Demographic information was collected at the time of PET MPI including age, sex, body mass index (BMI), family history of CAD, smoking status, hypertension, diabetes, and dyslipidemia. Medical history was determined through patient questionnaires and existing medical records.

### PET/CT MPI Imaging Protocol

PET studies were performed in accordance with the American Society of Nuclear Cardiology (ASNC) MPI guidelines^1^. A total of 25,407 patients (71.8%) underwent ^82^Rb PET-MPI, and 9,984 patients (28.2%) underwent ^13^NH_3_ PET-MPI. Pharmacologic stress agents included the vasodilators regadenoson (77.1%), dipyridamole (14.5%), or adenosine (7.3%) as well as dobutamine (1.1%). Reconstructed images were generated using vendor-recommended iterative reconstruction algorithms optimized for each scanner and site as previously reported ^8^.

### Image Quality Control and Quantification

All PET-MPI studies underwent centralized quality control by core laboratory technologists blinded to clinical data, as previously described ^8^. Image integrity, PET/CT registration, and motion were assessed, with manual corrections performed when necessary. Perfusion, flow, and functional parameters were quantified using QPET (Cedars-Sinai Medical Center, Los Angeles, CA).

### Perfusion and Myocardial Flow Reserve

Myocardial perfusion was assessed using automated quantitation of total perfusion deficit (TPD), which incorporates the extent and severity of perfusion abnormality. The magnitude of myocardial ischemia was quantified by ischemic TPD (stress TPD – rest TPD) [iTPD]. Rest and stress MBF with a 1-tissue compartment kinetic model were calculated by QPET ^9^. MFR was computed as the ratio of stress over the rest MBF. MFR was assessed both globally and regionally within the left anterior descending (LAD), left circumflex (LCX), and right coronary artery (RCA) territories according to the American Heart Association (AHA) 17-segment model.^10^. For separate per-vessel analysis, an average MBF and MFR in each vascular territory were used ^11^. Abnormal MFR was defined as global or regional MFR<2.0 ^1,12^. Abnormal stress MBF was defined as global or regional stress MBF <1.8 mL/min/g ^13,14^. Based on global and regional MFR, patients were categorized into three groups: MFR-Group 1 (normal global and regional MFR), MFR-Group 2 (normal global but abnormal regional MFR), and MFR-Group 3 (abnormal global and regional MFR). Patients were also categorized into three groups based on global and regional stress MBF. For MFR, group sizes were 19,744 (MFR-Group 1), 3,184 (MFR-Group 2), and 12,463 (MFR-Group 3), respectively; corresponding MBF group sizes were 21,572 (MBF-Group 1), 2,717 (MBF-Group 2), and 11,102 (MBF-Group 3) (Table 1 and Supplemental Table 1).

**Table 1.**
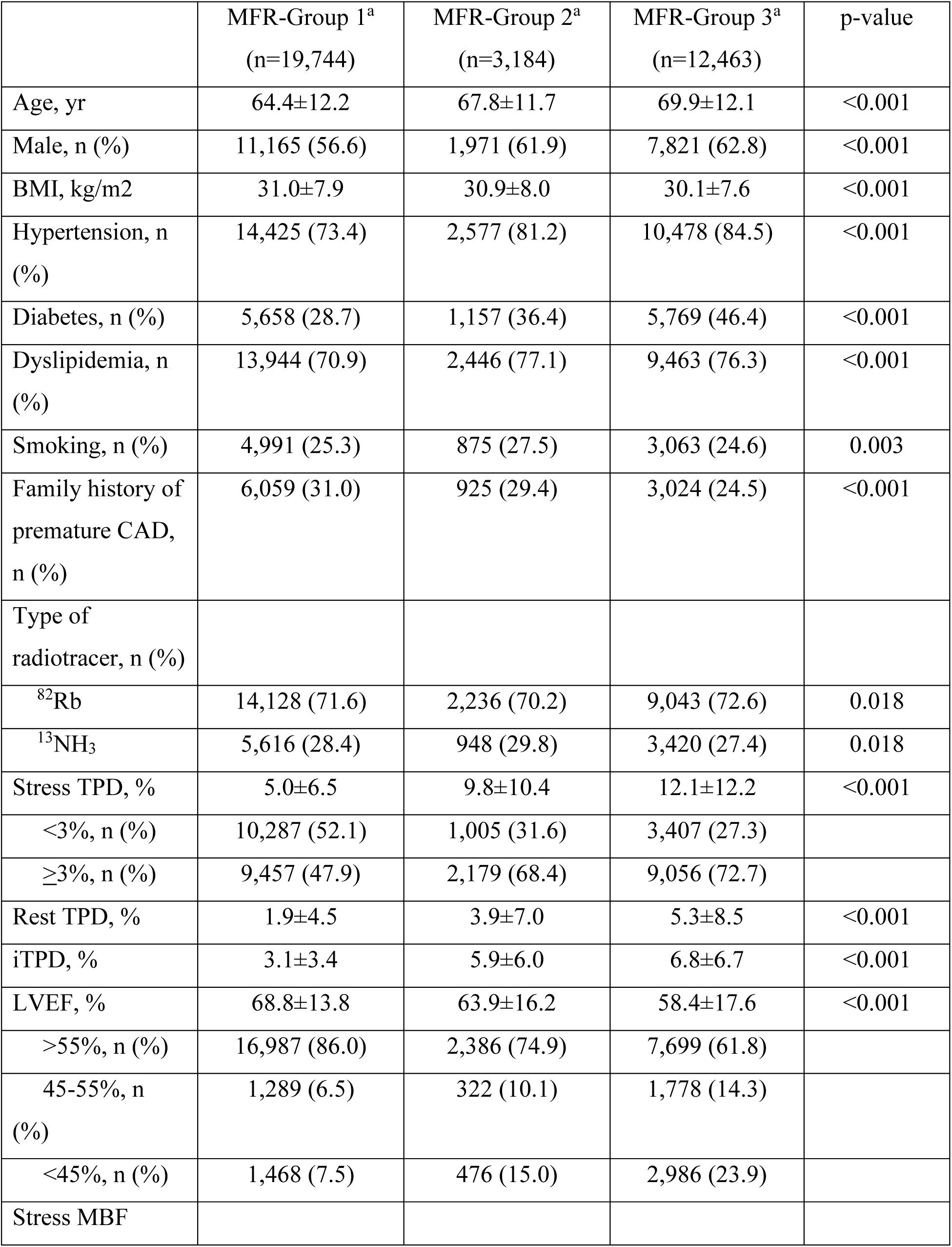
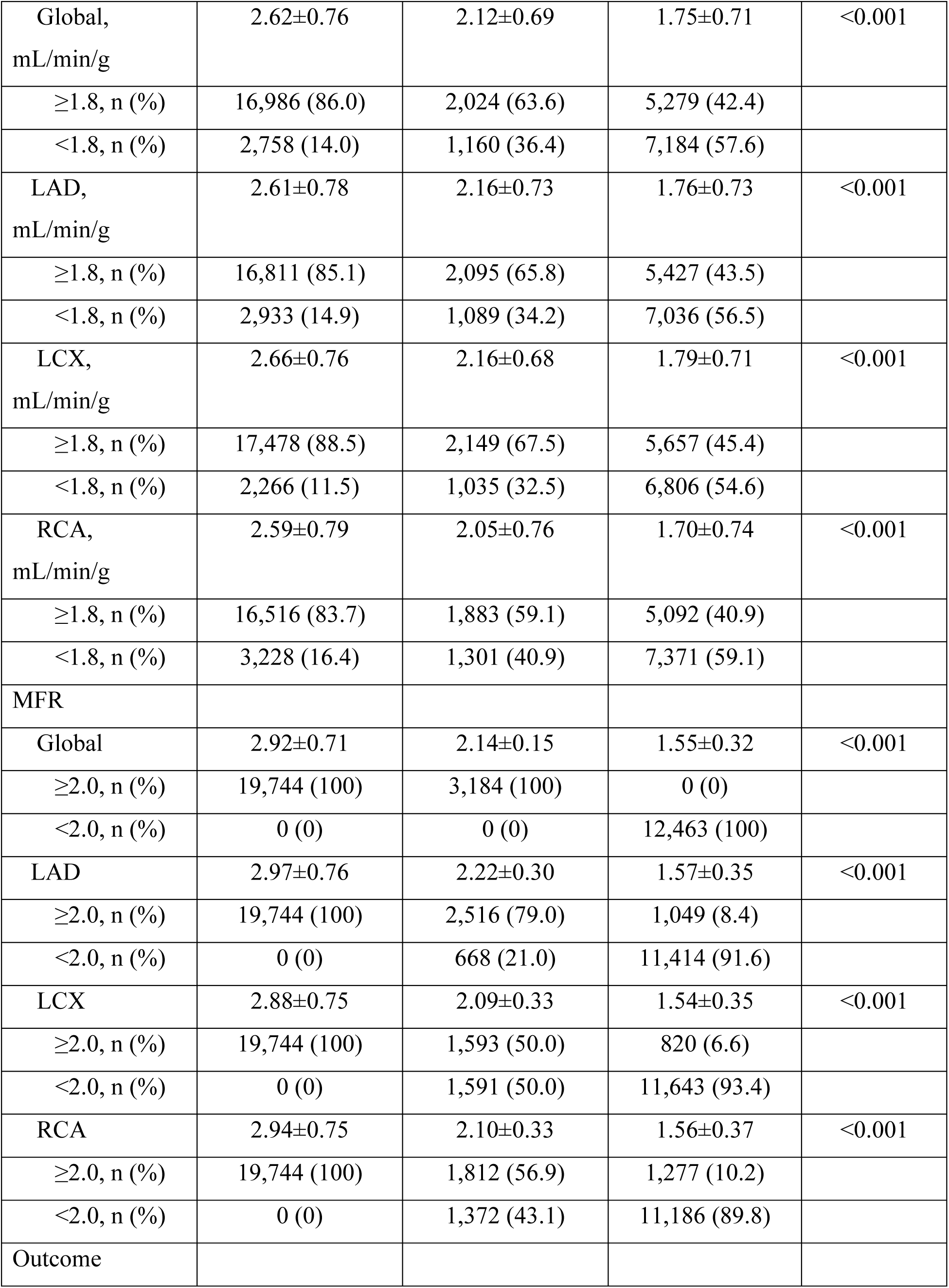
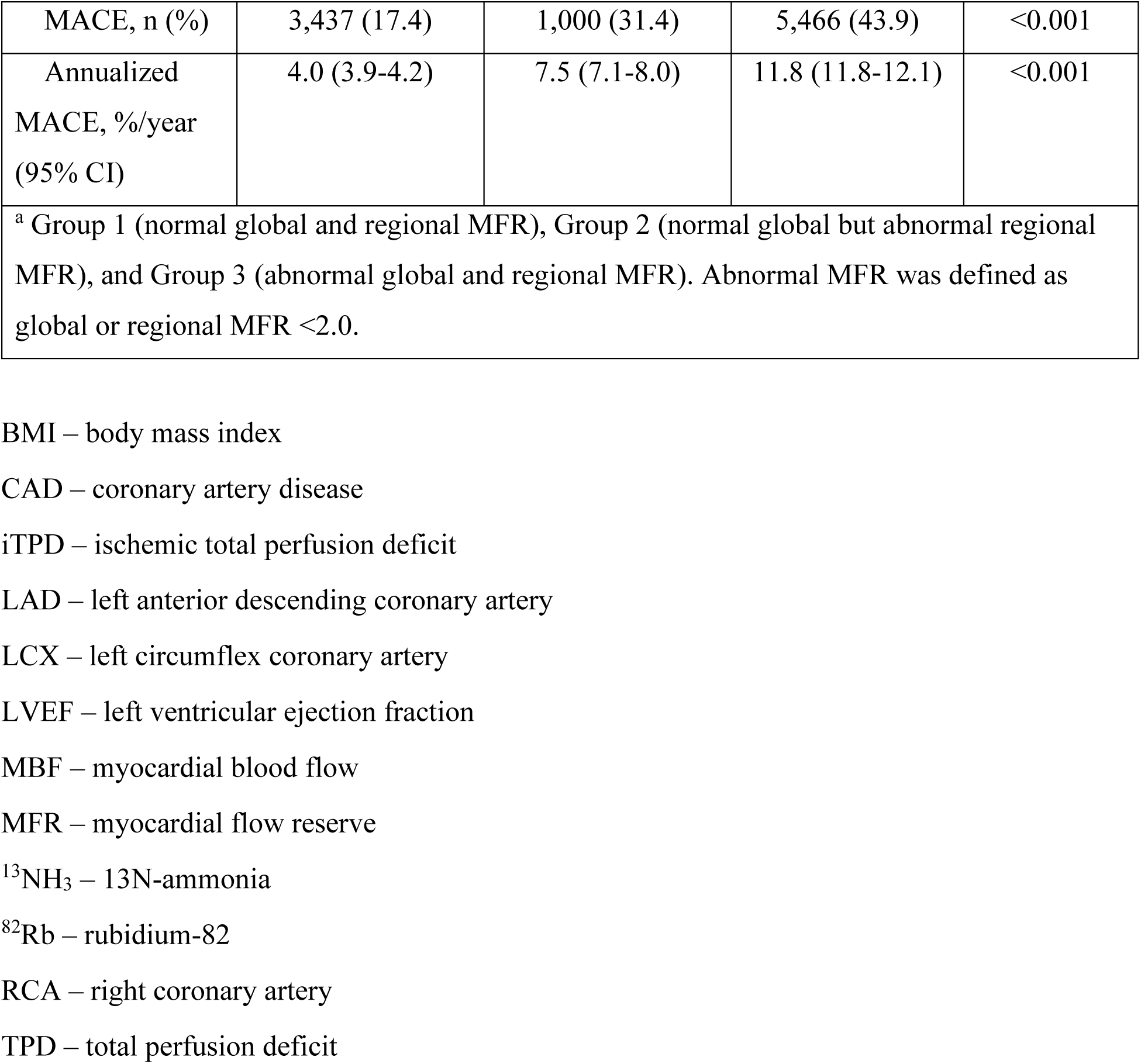
Baseline characteristics according to global and regional MFR.

### Outcomes

Our primary outcome was major adverse cardiovascular events (MACE) which included all-cause mortality, non-fatal myocardial infarction (MI), late revascularization (>90 days), and hospital admission for unstable angina (UA). Follow-up for clinical events was performed at each participating site. All events were reviewed and adjudicated by site investigators using standard clinical criteria ^8^.

### Statistical analysis

Continuous variables are presented as mean ± SD, and categorical variables are presented as counts (proportions). Continuous variables are compared using Student’s t-test or Mann-Whitney U test as appropriate. Categorical variables were compared using Pearson’s Chi-square test. Cumulative MACE incidence was assessed using the Kaplan–Meier method and compared with log-rank test. Cox proportional hazard regression analysis was used to calculate hazard ratio (HR) with 95% confidence interval (CI). Cox models were adjusted for age, sex, BMI, past medical history (hypertension, diabetes, dyslipidemia, smoking history), family history, stress TPD, and left ventricular ejection fraction (LVEF). Global chi-square analysis was used to assess the incremental value of regional MFR/stress MBF for MACE prediction using three models: (1) primary baseline model (age, sex, BMI, hypertension, diabetes, dyslipidemia, smoking, family history of premature CAD, LVEF, stress TPD), (2) primary baseline model + global MFR/MBF, and (3) primary baseline model + global MFR/MBF + regional MFR/MBF. Category-free net reclassification improvement (NRI) was calculated using Cox model-derived risk scores from the model incorporating global MFR/MBF and the model additionally incorporating regional MFR/MBF. The discriminatory performance of regional MFR and regional stress MBF for predicting MACE was evaluated using the area under the receiver operating characteristic curve (AUC), and comparisons were performed using the method described by DeLong et al ^15^. Missing covariate data were handled using complete-case analysis. Missing data were limited to BMI (n=201), hypertension (n=161), diabetes (n=41), dyslipidemia (n=161), smoking (n=15), family history (n=374), and stress LVEF (n=206). Of 35,391 patients, 34,920 had eligible survival-time observations, and 34,109 were included in the multivariable Cox analyses after excluding 811 patients with missing covariate data. Patients without MACE were censored at the last available follow-up. All statistical tests were two-sided. A p-value <0.05 was considered significant. Stata 17 was used for all analyses (StataCorp, College Station, TX, USA).

## Results

### Baseline Characteristics

In the overall study population, the mean age was 66.7 ± 12.4 years, with 59.2% (n=20,957) being male and a mean global MFR of 2.37. Baseline characteristics by MFR-based grouping are summarized in **Table 1**. Age, hypertension, and diabetes prevalence were lowest in MFR-Group 1 and highest in MFR-Group 3 (all *p* < 0.001). Stress TPD was highest in MFR-Group 3 (12.1% vs. 9.8% vs. 5.0%), with an elevated rest TPD (5.3% vs. 3.9% vs. 1.9%) and iTPD (6.8% vs. 5.9% vs. 3.1%) compared to Groups 2 and 1 (all *p* < 0.001). Global stress MBF was lowest in MFR-Group 3 (1.75 vs. 2.12 vs. 2.62 mL/min/g), with 57.6% exhibiting MBF <1.8 mL/min/g. Regional stress MBF <1.8 mL/min/g was predominant in MFR-Group 3 for LAD (56.5%), LCX (54.6%), and RCA (59.1%).

### Effect of Regional MFR on MACE

During a mean follow-up of 4.0 ± 2.9 years, 9,903 patients (28.0%) had MACE of the 35,391 patients in the entire cohort with an annualized MACE rate of 6.8 (6.7-7.0) %/year. MFR-Group 3 had the worst outcomes, followed by MFR-Group 2, while MFR-Group 1 had the best (11.8%/year vs. 7.5%/year vs. 4.0%/year, *p* < 0.001) (**Table 1**). Kaplan-Meier survival analysis showed that at 6 years, event-free survival was ∼80% in MFR-Group 1, ∼65% in MFR-Group 2, and ∼50% in MFR-Group 3 (**Figure 1**). Characteristics differences between the MACE group and non-MACE group are also shown in **Supplemental Table 2**. Global MFR was significantly lower in the MACE group (2.02 vs 2.50, *p* <0.001), with 55.2% of MACE group having an MFR <2.0. **Supplemental Table 2** shows that regional MFR <2.0 was predominant in the MACE group across the LAD (53.8%), LCX (56.0%) and RCA (55.3%) territories.

**Figure 1.**
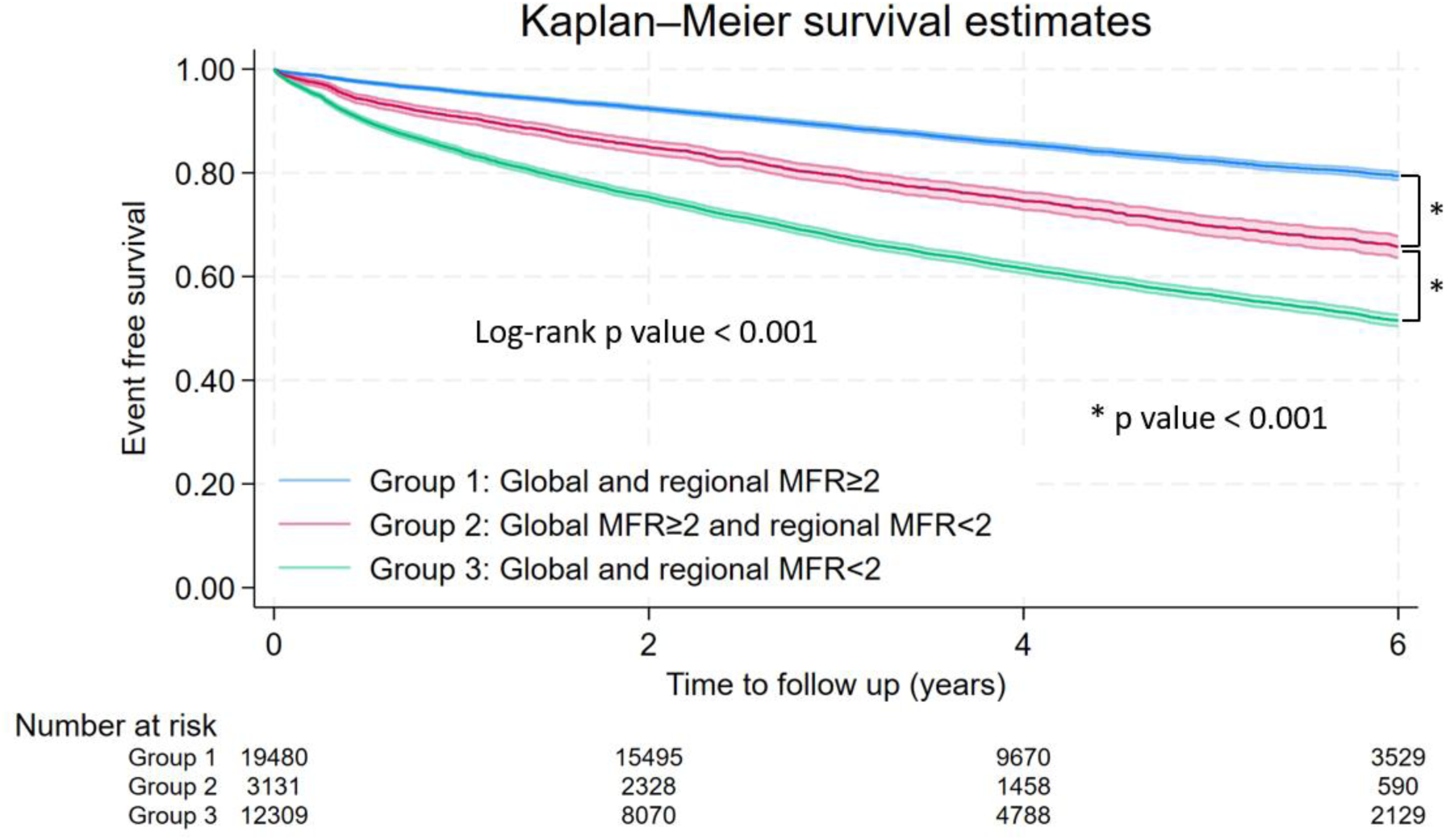
Kaplan–Meier curves for MACE according to global and regional MFR. Kaplan–Meier estimates of MACE across the 3 groups defined by global and regional MFR (log-rank *p*<0.001). MACE – major adverse cardiovascular event MFR – myocardial flow reserve

**Table 2** shows the univariable and multivariable Cox proportional hazards analyses to predict MACE. In the multivariable analysis, abnormal global MFR and abnormal regional MFR in any artery were independent predictors of MACE (both *p* < 0.05). For individual MACE components, abnormal regional MFR remained significantly associated with all-cause mortality, non-fatal MI, late revascularization, and unstable angina (**Supplemental Table 3**). Global chi-square analysis (**Table 3**) showed that addition of regional MFR to the primary baseline model incorporating global MFR significantly improved prediction of MACE (χ^2^ values: 4,871 vs 4,784, *p* < 0.001). AUC analysis using a primary baseline model showed that regional MFR significantly improved discrimination for MACE. The AUC increased from 0.710 to 0.729 with regional MFR (*p* < 0.001) (**Figure 2**).

**Table 2.**
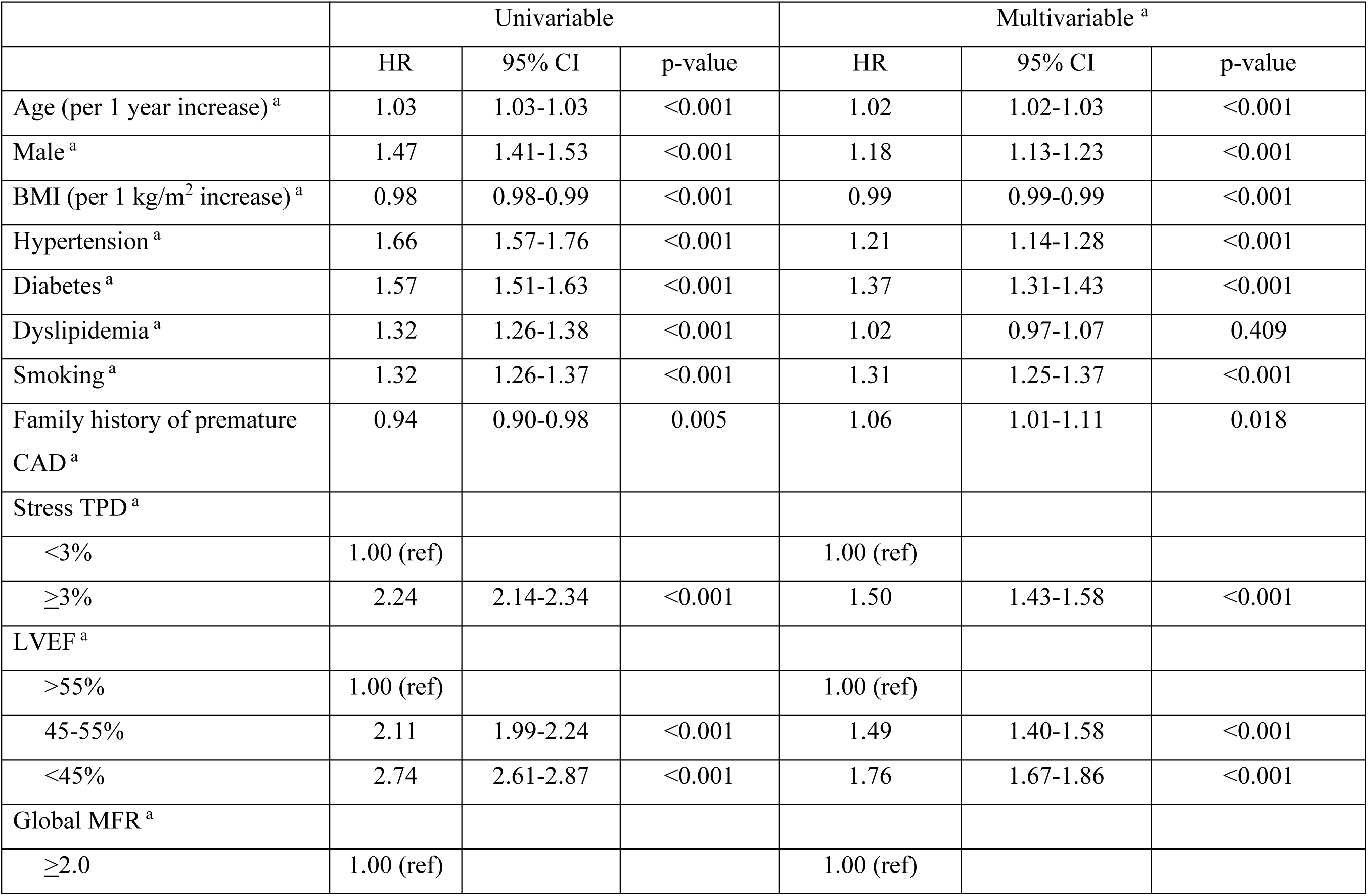
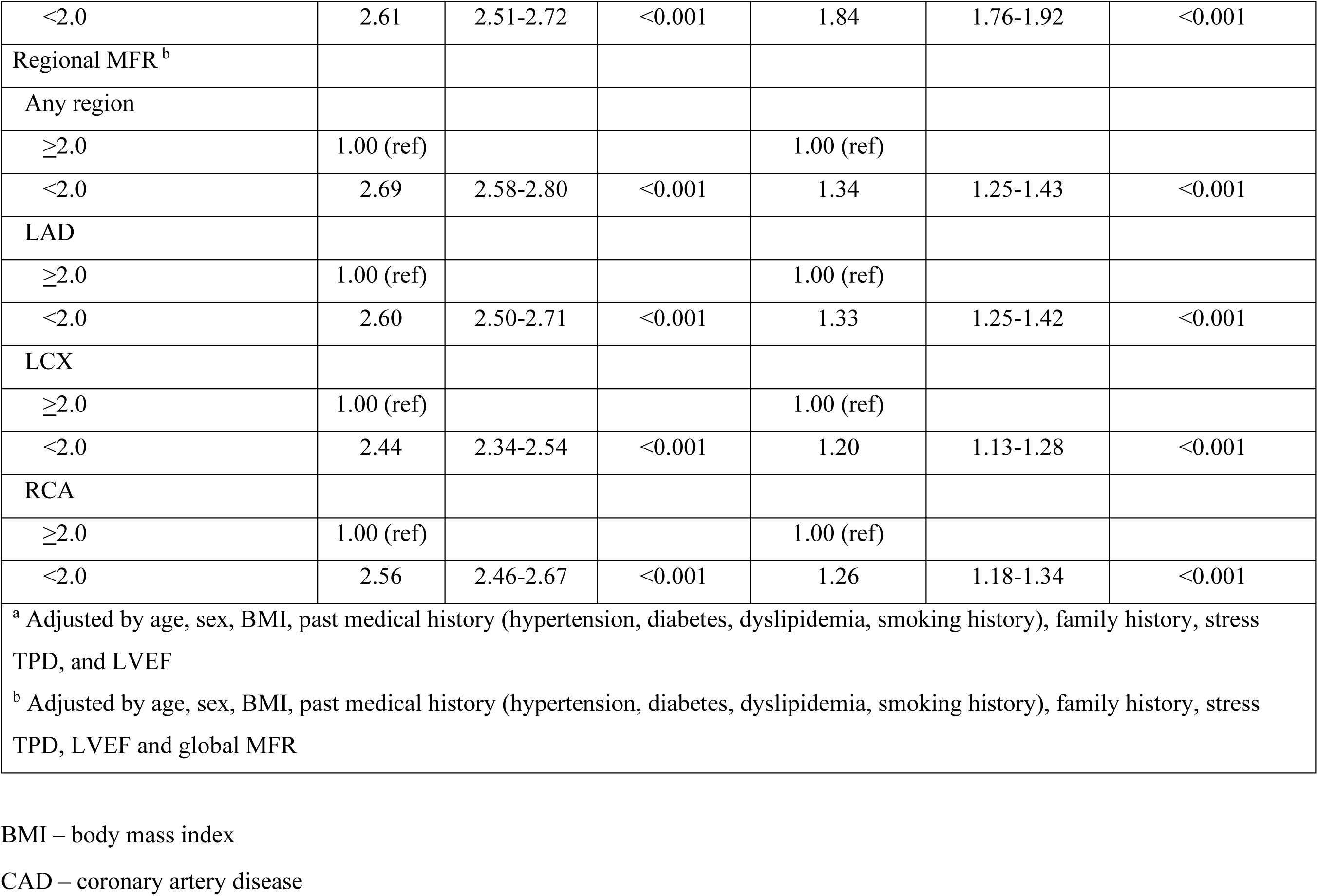
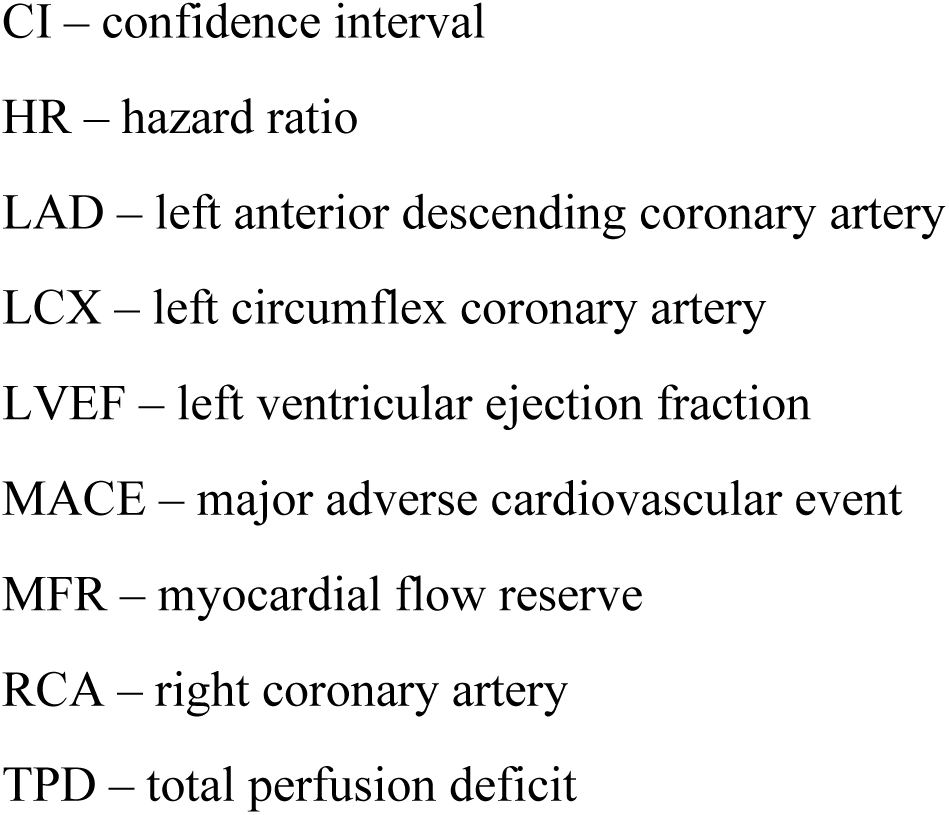
Cox regression analysis for prediction of MACE.

**Table 3.** Added benefit of regional MFR and MBF over conventional PET-MPI approach.

| Flow assessment | Model $\chi^2$ analysis | | | | | NRI analysis | | | |
| --- | --- | --- | --- | --- | --- | --- | --- | --- | --- |
| | Baseline model $\chi^2$ <sup>a</sup> | + Global flow | + Regional flow | $\Delta\chi^2$ with regional flow | p-value | NRI overall | % event | % non-event | p-value |
| Regional MFR <2.0 | 4,149 | 4,784 | 4,871 | 87 | <0.001 | 41.9% | 9.7% | 32.1% | <0.001 |
| Regional stress MBF <1.8 mL/min/g | 4,149 | 4,364 | 4,446 | 82 | <0.001 | 31.3% | -11.5% | 42.8% | <0.001 |
| <sup>a</sup> Primary baseline model included age, sex, BMI, hypertension, diabetes, dyslipidemia, current smoking status, family history of premature CAD, stress TPD and LVEF |  |  |  |  |  |  |  |  |  |

BMI – body mass index
CAD – coronary artery disease
LVEF – left ventricular ejection fraction
MBF – myocardial blood flow
MFR – myocardial flow reserve
MPI – myocardial perfusion imaging
NRI – net reclassification improvement
PET – positron emission tomography
TPD – total perfusion deficit

**Figure 2.**
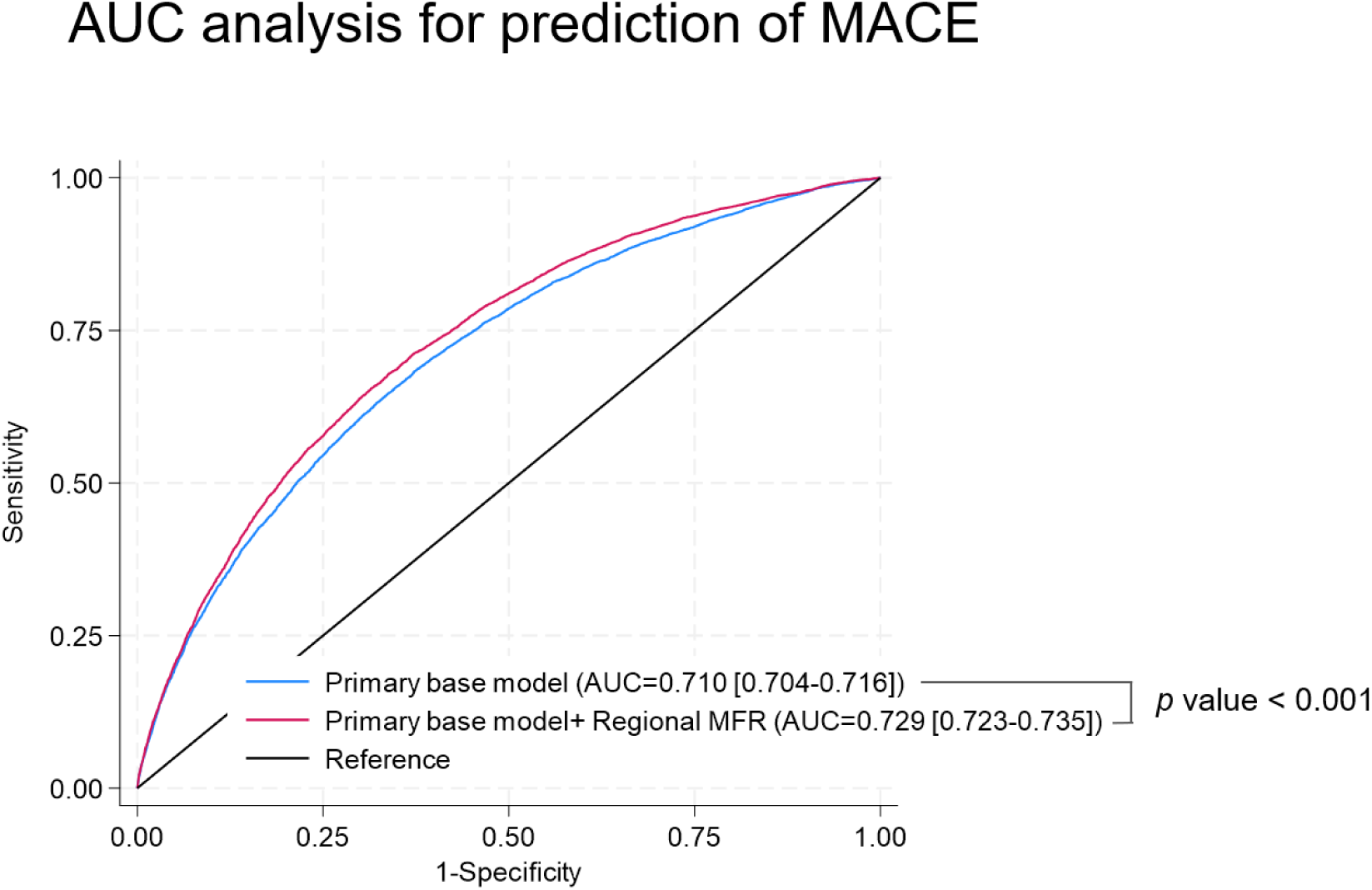
Incremental prognostic value of regional MFR. Receiver operating characteristic curves comparing the baseline model with and without regional MFR. Addition of regional MFR increased the area under the curve from 0.710 to 0.729 (*p* < 0.001). AUC – area under the receiver operating characteristics curve MACE – major adverse cardiovascular event MFR – myocardial flow reserve

The NRI was 41.9%, with a net increase of 9.7% for events and 32.1% for non-events. In subgroup analyses (**Figure 3**), abnormal regional MFR was associated with MACE across all subgroups. Significant interactions were observed for age, sex, stress TPD, LVEF, and radiotracer type (all *p* for interaction < 0.05), but not for diabetes. The association between abnormal regional MFR and MACE was stronger in patients aged <70 years, women, those with stress TPD <3%, and LVEF ≥45%. Importantly, abnormal regional MFR remained significantly associated with MACE in patients undergoing both ^82^Rb and ^13^NH_3_ PET imaging.

**Figure 3.**
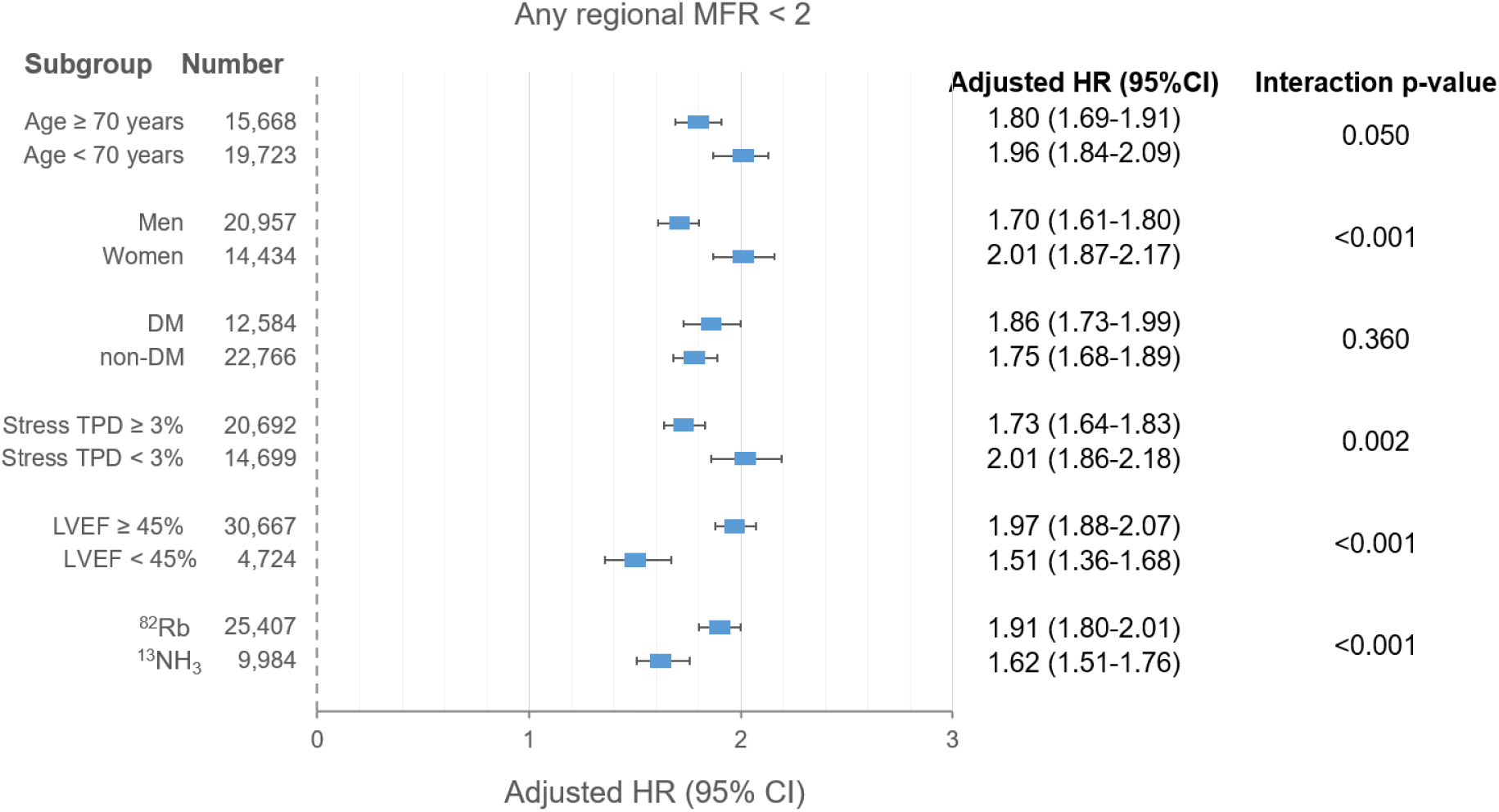
Subgroup analysis for independent prognostic value of regional MFR on MACE. Abnormal regional MFR was associated with MACE across all subgroups. Hazard ratios were adjusted for demographic characteristics, cardiovascular risk factors, stress TPD, and left ventricular ejection fraction. BMI – body mass index CI – confidence interval DM – diabetes mellitus HR – hazard ratio LVEF – left ventricular ejection fraction ^13^NH_3_ – 13N-ammonia MACE – major adverse cardiovascular event MFR – myocardial flow reserve ^82^Rb – rubidium-82 TPD – total perfusion deficit

### Risk stratification by regional MFR across conventional PET-MPI findings

To further evaluate the incremental prognostic value of regional MFR, annualized MACE rates were assessed according to stress TPD and global MFR categories (**Figure 4**). For this analysis, stress TPD ≥3% was defined as abnormal ^16^. Abnormal regional MFR was associated with significantly higher annualized MACE rates across all stress TPD categories, including patients with minimal ischemic burden (stress TPD <1%) (**Figure 4A**). Similarly, regional MFR significantly stratified risk among patients with preserved (≥2.5) and borderline (2.0 to <2.5) global MFR, although all patients with global MFR <2.0 had abnormal regional MFR by definition (**Figure 4B**). Regional MFR also provided significant risk stratification in both normal and abnormal stress TPD groups with preserved global MFR (**Figure 4C**). Notably, among patients with normal perfusion and preserved global MFR, annualized MACE rates were 2.8% and 5.0% in patients with normal and abnormal regional MFR, respectively (*p* < 0.001). **Supplemental Figure 1** shows representative case with preserved global but abnormal regional MFR who experienced MACE.

**Figure 4.**
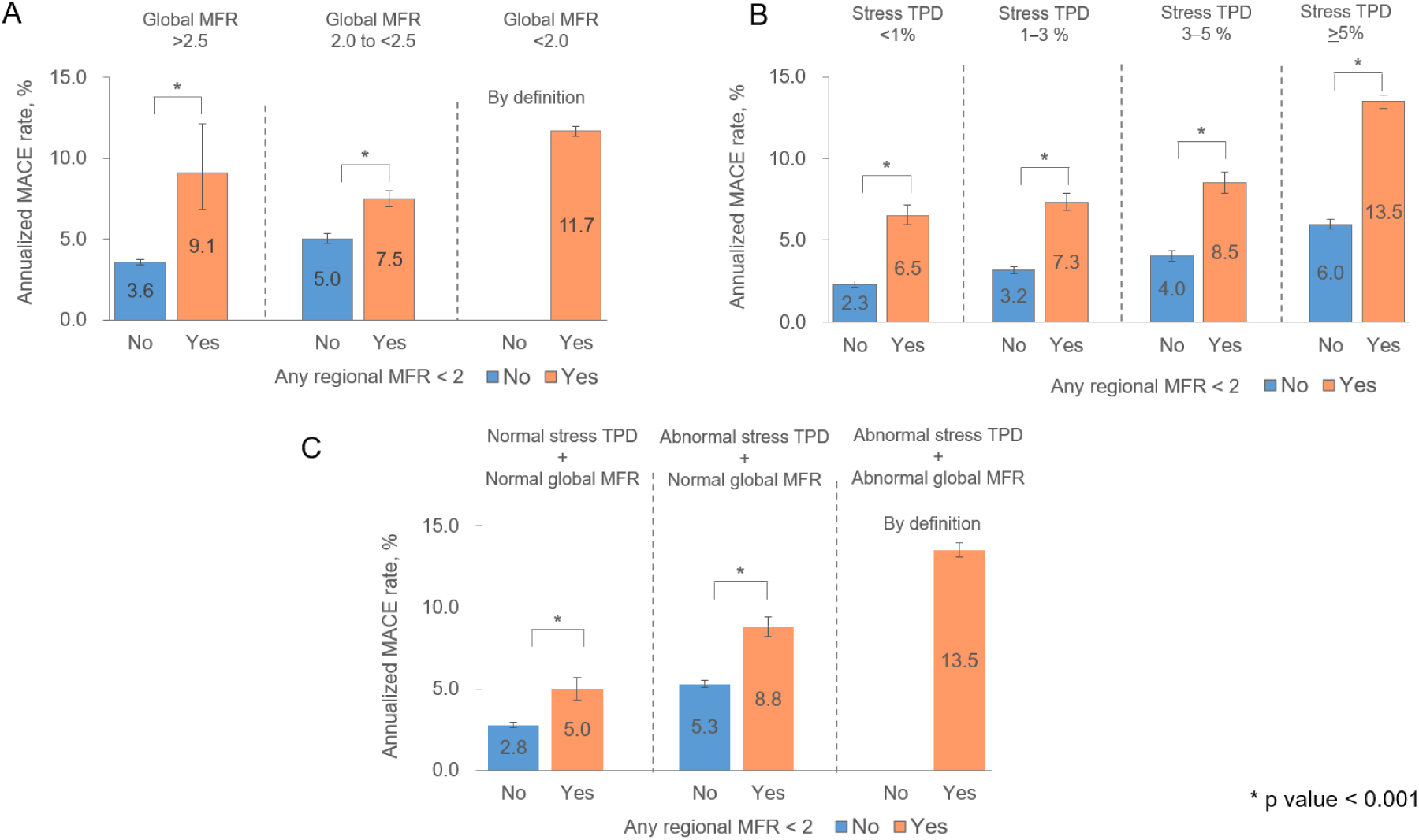
Risk stratification by regional MFR across stress TPD and global MFR categories. Annualized MACE rates according to regional MFR status, stratified by stress TPD and global MFR. All patients with global MFR <2.0 had abnormal regional MFR by definition. MACE – major adverse cardiovascular event MFR – myocardial flow reserve TPD – total perfusion deficit

### Effect of Regional Stress MBF on MACE

Baseline characteristics by MBF-based grouping are summarized in **Supplemental Table 1**. Overall, findings based on regional stress MBF were consistent with those observed for MFR, although the associations were numerically weaker. Global MFR was lowest in MBF-Group 3 (1.82 vs. 2.25 vs. 2.66), with 65.6% of patients having MFR <2.0. During follow-up, MBF-Group 3 had the highest event rate, followed by MBF-Group 2 and MBF-Group 1 (11.7%/year vs. 8.5%/year vs. 4.5%/year, *p* < 0.001) (**Supplemental Table 1** and **Supplemental Figure 2**). Multivariable analysis identified abnormal global MBF and any abnormal regional MBF as independent MACE predictors (all *p* <0.05) (**Supplemental Table 4**). Global chi-square analysis (**Table 3**) showed that adding regional MBF to conventional PET-MPI improved MACE prediction (χ^2^= 4,446 vs. 4,364, *p* < 0.001).

## Discussion

In this study, we demonstrate that abnormal regional MFR assessed by ^82^Rb and ^13^NH_3_ PET-MPI was associated with increased risk of MACE in patients with normal global MFR, independent of perfusion defects and conventional risk factors. Notably, incorporating regional MFR into a model including clinical risk factors and conventional MPI findings such as stress TPD, LVEF, and global MFR, improves both discriminatory and reclassification value for MACE prediction. Importantly, regional MFR further stratified risk among patients with otherwise reassuring PET-MPI findings, including those with preserved global MFR and minimal perfusion abnormalities. Finally, regional stress MBF was associated with MACE in patients with normal global stress MBF. These findings suggest that regional flow assessment can significantly enhance the utility of risk stratification of PET-MPI.

A few small-sized studies have explored the prognostic value of regional flow parameters. For instance, Bom et al. found that regional MFR abnormalities, as assessed by H_2_O PET and defined as two consecutive segments with the lowest values within a vascular territory, were associated with increased risk of MACE ^17^. In a more recent study, Felten et al. reported that reduced regional MFR within a vessel territory, assessed by ^13^NH_3_ PET in 150 patients, provided independent prognostic value ^18^. One previous study has evaluated the prognostic significance of regional MFR using ^82^Rb PET-MPI ^6^. In this study, 94 patients with normal global MFR but abnormal regional MFR (<1.5) were analyzed in a subgroup analysis, suggesting a potential association between abnormal regional MFR and MACE. In contrast, the present investigation leveraged a substantially larger cohort of more than 35,000 patients undergoing ^82^Rb and ^13^NH_3_ PET-MPI and applied a clinically relevant MFR threshold (<2.0), identifying 3,184 patients with preserved global MFR but impaired regional MFR in at least one vascular territory. Our findings demonstrate the independent and incremental prognostic value of regional MFR for predicting MACE beyond conventional PET-MPI findings and clinical risk factors. Moreover, subgroup analyses showed that the association between abnormal regional MFR and MACE was consistent across both ^82^Rb and ^13^NH_3_ PET-MPI tracers. Our study is the largest-scale investigation to date to explore the prognostic value of regional MFR in patients undergoing ^82^Rb and ^13^NH_3_ PET-MPI.

Most existing literature on MFR assessment using PET-MPI has focused on global MFR, which represents the average MFR across all myocardial segments, rather than regional MFR ^13,19,20^. Since early-stage focal CAD may subtly affect myocardial blood flow without causing significant perfusion defects, MFR measurements related to coronary distribution can offer valuable clinical insights ^7,21,22^. Although current guidelines recommend including both global and regional MFR measurements in routine reports, the prognostic evidence for regional MFR, particularly with ^82^Rb PET-MPI, remains limited ^1,5^. Our study shows that impaired regional MFR in any specific vessel territory (LAD, LCX, or RCA) is independently associated with a higher risk of MACE. This underscores the clinical significance of subtle MFR reductions, even in patients with normal global MFR, a group typically considered to have an excellent prognosis.

Interestingly, subgroup analyses suggested that the association between abnormal regional flow parameters and MACE was more pronounced in patients aged <70 years, women, and those with stress TPD <3% or LVEF ≥45%, groups generally considered to have lower baseline risk. This observation is particularly relevant given that, despite the relatively high-risk characteristics of the overall cohort, the majority of patients (64.8%) had preserved global MFR, and a substantial proportion (41.5%) had normal perfusion. In patients with substantial perfusion abnormalities or globally impaired MFR, elevated cardiovascular risk may already be apparent from conventional PET-MPI findings. In contrast, risk stratification remains challenging when global MFR is preserved and perfusion abnormalities are absent or minimal. Notably, even among patients with normal perfusion and preserved global MFR, abnormal regional MFR was associated with a substantially higher annualized MACE rate (5.0% vs. 2.8%). Regional MFR also provided significant risk stratification across all stress TPD categories, including patients with stress TPD <1%, and among those with preserved global MFR. These findings suggest that regional MFR may identify residual risk not captured by conventional perfusion assessment or global MFR and may therefore refine risk assessment in patients with otherwise reassuring PET-MPI findings.

In our study, the prognostic association of regional MFR appeared numerically stronger in the ^82^Rb cohort than in the ^13^NH_3_ cohort. Optimal MFR thresholds may differ across PET tracers due to differences in tracer kinetics and quantification characteristics ^23^. Applying the same cutoff to both tracers may therefore have resulted in relative under-detection of abnormal MFR in the ^13^NH_3_ cohort, potentially attenuating its observed prognostic association. These observations should be interpreted cautiously, and future studies using tracer-specific thresholds may help clarify these differences.

We also demonstrated that regional stress MBF was effective for risk stratification, even in patients with normal global stress MBF. While MFR is a well-established marker of coronary vascular function, stress MBF has also been associated with risk stratification, supporting our findings ^24,25^. Findings for regional stress MBF were generally consistent with those observed for regional MFR, with both regional measures providing significant incremental prognostic information beyond their corresponding global measures. Although the improvement in model fit was numerically greater for the MFR-based model, the incremental increases in χ² were relatively similar, as shown in **Table 3**. These results suggest that both regional MFR and regional stress MBF may offer complementary value for risk stratification in patients undergoing PET MPI.

## Limitations

This study has several limitations. First, as a retrospective observational study, selection bias and residual confounding cannot be excluded. Furthermore, the findings may not be generalizable to populations not referred for PET MPI. Second, we did not apply population-specific thresholds for flow assessment, and the optimal cutoff values may differ between regional and global MFR (e.g., segmental MFR, vessel MFR, and global MFR) ^7,26^. However, we followed the ASNC/Society of Nuclear Medicine and Molecular Imaging-recommended threshold for MFR and stress MBF, as established in prior studies, to ensure comparability ^1,12–14^. Third, different pharmacologic stressors may influence MBF and MFR values. In our cohort, regadenoson was most frequently used in MFR/MBF-Group 1 (**Supplemental Table 5**) (*p* <0.001 for trend). On the other hand, after additional adjustment for stressor agent in the multivariable Cox model based on the primary baseline model, regional MFR <2.0 and stress MBF <1.8 mL/min/g remained significantly associated with MACE (**Supplemental Table 6**). Fourth, we did not assess the correlation between regional MFR and coronary territories identified by invasive coronary angiography in individual patients, making the relationship between coronary events and specific territories unclear ^27^. Further research is needed for validation.

## Conclusion

Abnormal regional MFR provided incremental prognostic value beyond conventional PET-MPI findings, including global MFR. Regional MFR may refine risk stratification, particularly in patients with preserved global MFR and minimal perfusion abnormalities.

## Abbreviations List

CAD: coronary artery disease
MACE: major adverse cardiovascular event
MBF: myocardial blood flow
MFR: myocardial flow reserve
MPI: myocardial perfusion imaging
13NH3: ^13^N-ammonia
PET: positron emission tomography
82Rb: rubidium-82
TPD: total perfusion deficit

## Acknowledgement

We would like to thank all participating institutions and investigators who contributed clinical and imaging data to the REFINE PET registry. We are especially grateful to the data managers, research coordinators, and technical staff at each site for their critical roles in data collection. We also acknowledge the support of the core laboratory at Cedars-Sinai Medical Center for data harmonization, imaging analysis, and infrastructure support. Special thanks are extended to the imaging technologists, software engineers, and data scientists who developed and implemented the automated artificial intelligence tools used in this study. This work was supported in part by grants R35HL161195 from the National Heart, Lung, and Blood Institute and R01EB034586 from the National Institute of Biomedical Imaging and Bioengineering of the National Institutes of Health.

