## Supplemental Material for "Incremental Prognostic Value of Regional Myocardial Flow Reserve Beyond Global Flow and Perfusion: REFINE PET"

P2-4: Supplemental Table 1
P5-7: Supplemental Table 2
P8-9: Supplemental Table 3
P10-11: Supplemental Table 4
P12: Supplemental Table 5
P13: Supplemental Table 6
P14: Supplemental Figure 1
P15: Supplemental Figure 2

**Supplemental Table 1. Baseline characteristics according to global and regional stress MBF**

| | MBF-Group 1[a] (n=21,572) | MBF-Group 2[a] (n=2,717) | MBF-Group 3[a] (n=11,102) | p-value |
|---|---|---|---|---|
| Age, yr | 65.9±12.5 | 67.7±12.4 | 68.0±12.0 | <0.001 |
| Male, n (%) | 10,370 (48.1) | 1,904 (70.1) | 8,683 (78.2) | <0.001 |
| BMI, kg/$m^2$ | 30.2±7.5 | 30.7±7.8 | 31.5±8.3 | <0.001 |
| Hypertension, n (%) | 16,141 (75.2) | 2,155 (79.7) | 9,184 (82.9) | <0.001 |
| Diabetes, n (%) | 6,405 (29.7) | 1,051 (38.7) | 5,128 (46.3) | <0.001 |
| Dyslipidemia, n (%) | 15,179 (70.8) | 2,043 (75.6) | 8,631 (77.9) | <0.001 |
| Smoking, n (%) | 5,071 (23.5) | 744 (27.4) | 3,114 (28.1) | <0.001 |
| Family history of premature CAD, n (%) | 6,299 (29.6) | 735 (27.3) | 2,974 (27.0) | <0.001 |
| Type of radiotracer, n (%) | | | | |
| $^{82}Rb$ | 17,195 (79.7) | 1,768 (65.1) | 6,444 (58.0) | <0.001 |
| $^{13}NH_3$ | 4,377 (20.3) | 949 (34.9) | 4,658 (42.0) | <0.001 |
| Stress TPD, % | 4.1±5.1 | 10.3±9.1 | 14.7±12.8 | <0.001 |
| <3%, n (%) | 12,212 (56.6) | 618 (22.7) | 1,869 (16.8) | |
| ≥3%, n (%) | 9,360 (43.4) | 2,099 (77.3) | 9,233 (83.2) | |
| Rest TPD, % | 1.3±2.9 | 3.9±6.2 | 7.1±9.5 | <0.001 |
| iTPD, % | 2.9±3.3 | 6.4±5.7 | 7.7±6.8 | <0.001 |
| LVEF, % | 70.8±12.3 | 62.2±14.1 | 53.5±17.0 | <0.001 |
| >55%, n (%) | 19,386 (89.9) | 2,011 (74.0) | 5,675 (51.1) | <0.001 |
| 45-55%, n (%) | 1,186 (5.5) | 334 (12.3) | 1,869 (16.8) | |
| <45%, n (%) | 1,000 (4.6) | 372 (13.7) | 3,558 (32.1) | |
| Stress MBF | | | | |
| Global, mL/min/g | 2.78±0.62 | 1.94±0.13 | 1.34±0.32 | <0.001 |
| ≥1.8, n (%) | 21,572 (100.0) | 2,717 (100.0) | 0 (0.0) | |
| <1.8, n (%) | 0 (0.0) | 0 (0.0) | 11,102 (100.0) | |
| LAD, mL/min/g | 2.78±0.65 | 1.98±0.26 | 1.34±0.36 | <0.001 |
| ≥1.8, n (%) | 21,572 (100.0) | 1,968 (72.4) | 793 (7.1) | |

| | | | | |
|---|---|---|---|---|
| <1.8, n (%) | 0 (0.0) | 749 (27.6) | 10,309 (92.9) | |
| LCX, mL/min/g | 2.80±0.65 | 2.01±0.31 | 1.43±0.38 | <0.001 |
| ≥1.8, n (%) | 21,572 (100.0) | 1,952 (71.8) | 1,760 (15.9) | |
| <1.8, n (%) | 0 (0.0) | 765 (28.1) | 9,342 (84.1) | |
| RCA, mL/min/g | 2.76±0.64 | 1.81±0.28 | 1.29±0.38 | <0.001 |
| ≥1.8, n (%) | 21,572 (100.0) | 1,145 (42.1) | 774 (7.0) | |
| <1.8, n (%) | 0 (0.0) | 1,572 (57.9) | 10,328 (93.0) | |
| MFR | | | | |
| Global | 2.66±0.82 | 2.25±0.70 | 1.82±0.64 | <0.001 |
| ≥2.0, n (%) | 17,388 (80.6) | 1,622 (59.7) | 3,918 (35.3) | |
| <2.0, n (%) | 4,184 (19.4) | 1,095 (40.3) | 7,184 (64.7) | |
| LAD | 2.70±0.86 | 2.33±0.75 | 1.86±0.69 | <0.001 |
| ≥2.0, n (%) | 17,486 (81.1) | 1,696 (62.4) | 4,127 (37.1) | |
| <2.0, n (%) | 4,086 (18.9) | 1,021 (37.6) | 6,975 (62.8) | |
| LCX | 2.62±0.85 | 2.23±0.74 | 1.82±0.68 | <0.001 |
| ≥2.0, n (%) | 16,688 (77.4) | 1,549 (57.0) | 3,920 (35.3) | |
| <2.0, n (%) | 4,884 (22.6) | 1,168 (43.0) | 7,182 (64.7) | |
| RCA | 2.70±0.85 | 2.22±0.73 | 1.81±0.68 | <0.001 |
| ≥2.0, n (%) | 17,487 (81.1) | 1,522 (56.0) | 3,824 (34.4) | |
| <2.0, n (%) | 4,085 (18.9) | 1,195 (44.0) | 7,278 (65.6) | |
| Outcome | | | | |
| MACE, n (%) | 4,177 (19.4) | 930 (34.2) | 4,796 (43.2) | <0.001 |
| Annualized MACE, %/year (95% CI) | 4.5 (4.4-4.7) | 8.5 (8.0-9.1) | 11.7 (11.4-12.1) | <0.001 |

[a] Group 1 (normal global and regional stress MBF), Group 2 (normal global but abnormal regional stress MBF), and Group 3 (abnormal global and regional stress MBF). Abnormal stress MBF was defined as global or regional stress MBF <1.8 ml/min/g.

BMI – body mass index
CAD – coronary artery disease
iTPD – ischemic total perfusion deficit

LAD – left anterior descending coronary artery
LCX – left circumflex coronary artery
LVEF – left ventricular ejection fraction
MACE – major adverse cardiovascular event
MBF – myocardial blood flow
MFR – myocardial flow reserve
$^{13}NH_3$ – $^{13}N$-ammonia
$^{82}Rb$ – rubidium-82
RCA – right coronary artery
TPD – total perfusion deficit

**Supplemental Table 2. Baseline characteristics according to MACE**

| | All (n=35,391) | MACE (n=9,903) | No MACE (n=25,488) | p-value |
|---|---|---|---|---|
| Age, yr | 66.7±12.4 | 69.6±11.9 | 65.5±12.4 | <0.001 |
| Male, n (%) | 20,957 (59.2) | 6,495 (65.6) | 14,462 (56.7) | <0.001 |
| BMI, kg/m$^2$ | 30.6±7.8 | 30.9±7.9 | 29.8±7.5 | <0.001 |
| Hypertension, n (%) | 27,480 (78.0) | 8,324 (84.7) | 19,156 (75.4) | <0.001 |
| Diabetes, n (%) | 12,584 (35.6) | 4,333 (43.8) | 8,251 (32.4) | <0.001 |
| Dyslipidemia, n (%) | 25,853 (73.4) | 7,505 (76.4) | 18,348 (72.2) | <0.001 |
| Smoking, n (%) | 8,929 (25.2) | 3,000 (30.3) | 5,929 (23.3) | <0.001 |
| Family history of premature CAD, n (%) | 10,008 (28.6) | 2,711 (27.8) | 7,297 (28.9) | 0.041 |
| Type of radiotracer, n (%) | | | | |
| 82Rb | 25,407 (71.8) | 6,617 (66.8) | 18,790 (73.7) | <0.001 |
| 13NH3 | 9,984 (28.2) | 3,286 (33.2) | 6,698 (26.3) | <0.001 |
| Stress TPD, % | 7.9±9.9 | 11.9±11.7 | 6.4±8.5 | <0.001 |
| <3%, n (%) | 14,699 (41.5) | 2,466 (24.9) | 12,233 (48.0) | |
| ≥3%, n (%) | 20,692 (58.5) | 7,437 (75.1) | 13,255 (52.0) | |
| Rest TPD, % | 3.3±6.6 | 5.4±8.4 | 2.5±5.6 | <0.001 |
| iTPD, % | 4.7±5.3 | 6.6±6.2 | 3.9±4.8 | <0.001 |
| LVEF, % | 64.7±16.2 | 57.7±17.5 | 67.5±14.8 | <0.001 |
| >55%, n (%) | 27,072 (76.5) | 6,001 (60.6) | 21,071 (82.7) | |
| 45-55%, n (%) | 3,389 (9.6) | 1,414 (14.3) | 1,975 (7.7) | |
| <45%, n (%) | 4,930 (13.9) | 2,488 (25.1) | 2,442 (9.6) | |
| Stress MBF | | | | |
| Global, mL/min/g | 2.27±0.84 | 1.94±0.82 | 2.39±0.81 | <0.001 |
| ≥1.8, n (%) | 24,289 (68.6) | 5,107 (51.6) | 19,182 (75.3) | |
| <1.8, n (%) | 11,102 (31.4) | 4,796 (48.4) | 6,306 (24.7) | |

| | | | | |
|---|---|---|---|---|
| LAD, mL/min/g | 2.27±0.86 | 1.95±0.83 | 2.39±0.83 | <0.001 |
| ≥1.8, n (%) | 24,333 (68.8) | 5,185 (52.4) | 19,148 (75.1) | |
| <1.8, n (%) | 11,058 (31.2) | 4,718 (47.6) | 6,340 (24.9) | |
| LCX, mL/min/g | 2.31±0.84 | 2.01±0.84 | 2.43±0.81 | <0.001 |
| ≥1.8, n (%) | 25,284 (71.4) | 5,533 (55.9) | 19,751 (77.5) | |
| <1.8, n (%) | 10,107 (28.6) | 4,370 (44.1) | 5,737 (22.5) | |
| RCA, mL/min/g | 2.23±0.87 | 1.87±0.85 | 2.36±0.85 | <0.001 |
| ≥1.8, n (%) | 23,491 (66.4) | 4,817 (48.6) | 18,674 (73.3) | |
| <1.8, n (%) | 11,900 (33.6) | 5,086 (51.4) | 6,814 (26.7) | |
| MFR | | | | |
| Global | 2.37±0.85 | 2.02±0.79 | 2.50±0.84 | <0.001 |
| ≥2.0, n (%) | 22,928 (64.8) | 4,437 (44.8) | 18,491 (72.6) | |
| <2.0, n (%) | 12,463 (35.2) | 5,466 (55.2) | 6,997 (27.5) | |
| LAD | 2.41±0.89 | 2.06±0.82 | 2.55±0.88 | <0.001 |
| ≥2.0, n (%) | 23,309 (65.9) | 4,576 (46.2) | 18,733 (73.5) | |
| <2.0, n (%) | 12,082 (34.1) | 5,327 (53.8) | 6,755 (26.5) | |
| LCX | 2.34±0.87 | 2.01±0.81 | 2.46±0.86 | <0.001 |
| ≥2.0, n (%) | 22,157 (62.6) | 4,359 (44.0) | 17,798 (69.8) | |
| <2.0, n (%) | 13,234 (37.4) | 5,544 (56.0) | 7,690 (30.2) | |
| RCA | 2.38±0.89 | 2.03±0.84 | 2.52±0.87 | <0.001 |
| ≥2.0, n (%) | 22,833 (64.5) | 4,428 (44.7) | 18,405 (72.2) | |
| <2.0, n (%) | 12,558 (35.5) | 5,475 (55.3) | 7,083 (27.8) | |

BMI – body mass index
CAD – coronary artery disease
iTPD – ischemic total perfusion deficit

LAD – left anterior descending coronary artery
LCX – left circumflex coronary artery
LVEF – left ventricular ejection fraction
MBF – myocardial blood flow
MFR – myocardial flow reserve
$^{13}NH_3$ – 13 N-ammonia
$^{82}Rb$ – rubidium-82

RCA – right coronary artery
TPD – total perfusion deficit

**Supplemental Table 3. HRs for Outcomes in Patients With abnormal regional MFR/ MBF vs Without abnormal regional MFR/MBF**

| | Any regional MFR | | Any regional stress MBF | |
|---|---|---|---|---|
| | ≥2.0 (n=15,647) | <2.0 (n=19,744) | ≥1.8, mL/min/g (n=21,572) | <1.8, mL/min/g (n=13,819) |
| All-cause mortality No. of events (%) | 2,077 (10.5) | 4,761 (30.4) | 2,995 (13.9) | 3,843 (27.8) |
| Unadjusted HR (95%CI) | 1.00 (ref) | 3.04 (2.88-3.20) [b] | 1.00 (ref) | 2.11 (2.01-2.21) [b] |
| Adjusted HR (95%CI) [a] | 1.00 (ref) | 1.96 (1.85-2.08) [b] | 1.00 (ref) | 1.31 (1.24-1.40) [b] |
| Non-fatal MI No. of events (%) | 669 (3.4) | 1,184 (7.6) | 573 (2.7) | 1,280 (9.3) |
| Unadjusted HR (95%CI) | 1.00 (ref) | 2.43 (2.20-2.68) [b] | 1.00 (ref) | 3.61 (3.26-3.99) [b] |
| Adjusted HR (95%CI) [a] | 1.00 (ref) | 1.55 (1.39-1.73) [b] | 1.00 (ref) | 2.18 (1.93-2.47) [b] |
| Late revascularization No. of events (%) | 947 (4.8) | 1,550 (9.9) | 914 (4.2) | 1,583 (11.5) |
| Unadjusted HR (95%CI) | 1.00 (ref) | 2.28 (2.11-2.47) [b] | 1.00 (ref) | 2.97 (2.74-3.23) [b] |
| Adjusted HR (95%CI) [a] | 1.00 (ref) | 1.67 (1.53-1.83) [b] | 1.00 (ref) | 1.98 (1.79-2.18) [b] |
| Unstable angina No. of events (%) | 518 (3.4) | 581 (5.2) | 416 (2.5) | 683 (7.0) |
| Unadjusted HR (95%CI) | 1.00 (ref) | 1.61 (1.41-1.82) [b] | 1.00 (ref) | 2.65 (2.33-3.02) [b] |
| Adjusted HR (95%CI) [a] | 1.00 (ref) | 1.17 (1.02-1.35) [b] | 1.00 (ref) | 1.90 (1.62-2.22) [b] |

[a] Adjusted by age, sex, BMI, hypertension, diabetes, dyslipidemia, current smoking status, family history of premature CAD, stress TPD and LVEF

[b] $p<0.001$

BMI – body mass index
CI – confidence interval
HR – hazard ratio
LVEF – left ventricular ejection fraction
MBF – myocardial blood flow
MFR – myocardial flow reserve
MI – myocardial infarction
TPD – total perfusion deficit

**Supplemental Table 4. Cox regression analysis for prediction of MACE**

| | Univariable | | | Multivariable [a] | | |
|---|---|---|---|---|---|---|
| | HR | 95% CI | p-value | HR | 95% CI | p-value |
| Global stress MBF, mL/min/g [a] | | | | | | |
| ≥1.8 | 1.00 (ref) | | | 1.00 (ref) | | |
| <1.8 | 2.36 | 2.27-2.46 | <0.001 | 1.55 | 1.48-1.63 | <0.001 |
| Regional stress MBF, mL/min/g [b] | | | | | | |
| Any region | | | | | | |
| ≥1.8 | 1.00 (ref) | | | 1.00 (ref) | | |
| <1.8 | 2.43 | 2.33-2.53 | <0.001 | 1.22 | 1.14-1.31 | <0.001 |
| LAD | | | | | | |
| ≥1.8 | 1.00 (ref) | | | 1.00 (ref) | | |
| <1.8 | 2.28 | 2.19-2.37 | <0.001 | 1.12 | 1.05-1.20 | <0.001 |
| LCX | | | | | | |
| ≥1.8 | 1.00 (ref) | | | 1.00 (ref) | | |
| <1.8 | 2.32 | 2.22-2.41 | <0.001 | 1.18 | 1.11-1.25 | <0.001 |
| RCA | | | | | | |
| ≥1.8 | 1.00 (ref) | | | 1.00 (ref) | | |
| <1.8 | 2.35 | 2.26-2.45 | <0.001 | 1.16 | 1.08-1.23 | <0.001 |

[a] Adjusted by age, sex, BMI, past medical history (hypertension, diabetes, dyslipidemia, smoking history), family history, stress TPD, and LVEF

[b] Adjusted by age, sex, BMI, past medical history (hypertension, diabetes, dyslipidemia, smoking history), family history, stress TPD, LVEF and global MFR

BMI – body mass index
CI – confidence interval

HR – hazard ratio
LAD – left anterior descending coronary artery
LCX – left circumflex coronary artery
LVEF – left ventricular ejection fraction
MACE – major adverse cardiovascular event
MBF – myocardial blood flow
RCA – right coronary artery
TPD – total perfusion deficit

**Supplemental Table 5. Stressor agent according to global and regional MFR/MBF**

| | MFR-Group 1[a] (n=19,744) | MFR-Group 2[a] (n=3,184) | MFR-Group 3[a] (n=12,463) | p-value |
|---|---|---|---|---|
| Stressor agent | | | | <0.001 |
| Dipyridamole | 2,569 (13.0) | 439 (13.8) | 2,107 (16.9) | |
| Adenosine | 1,074 (5.4) | 251 (7.9) | 1,257 (10.1) | |
| Regadenoson | 15,841 (80.2) | 2,464 (77.4) | 8,981 (72.1) | |
| Dobutamine | 245 (1.2) | 26 (0.8) | 113 (0.9) | |
| | MBF-Group 1[b] (n=21,572) | MBF-Group 2[b] (n=2,717) | MBF-Group 3[b] (n=11,102) | p-value |
| Stressor agent | | | | <0.001 |
| Dipyridamole | 3,036 (14.1) | 392 (14.4) | 1,687 (14.5) | |
| Adenosine | 958 (4.4) | 233 (8.6) | 1,391 (12.5) | |
| Regadenoson | 17,323 (80.3) | 2,069 (76.2) | 7,894 (71.1) | |
| Dobutamine | 247 (1.2) | 21 (0.8) | 116 (1.0) | |

[a] Group 1 (normal global and regional MFR), Group 2 (normal global but abnormal regional MFR), and Group 3 (abnormal global and regional MFR). Abnormal MFR was defined as global or regional MFR<2.0.

MBF – myocardial blood flow
MFR – myocardial flow reserve

**Supplemental Table 6. Cox regression analysis for prediction of MACE**

| | Multivariable [a] | | |
|---|---|---|---|
| | HR | 95% CI | p-value |
| Regional MFR [a] | | | |
| Any region | | | |
| $\geq$2.0 | 1.00 (ref) | | |
| <2.0 | 1.83 | 1.75-1.92 | <0.001 |
| Regional stress MBF, mL/min/g [a] | | | |
| Any region | | | |
| $\geq$1.8 | 1.00 (ref) | | |
| <1.8 | 1.55 | 1.47-1.63 | <0.001 |
| [a] Adjusted by age, sex, BMI, past medical history (hypertension, diabetes, dyslipidemia, smoking history), family history, stress TPD, LVEF and stressor agent | | | |

BMI – body mass index
CI – confidence interval
HR – hazard ratio
LVEF – left ventricular ejection fraction
MBF – myocardial blood flow
MFR – myocardial flow reserve
TPD – total perfusion deficit

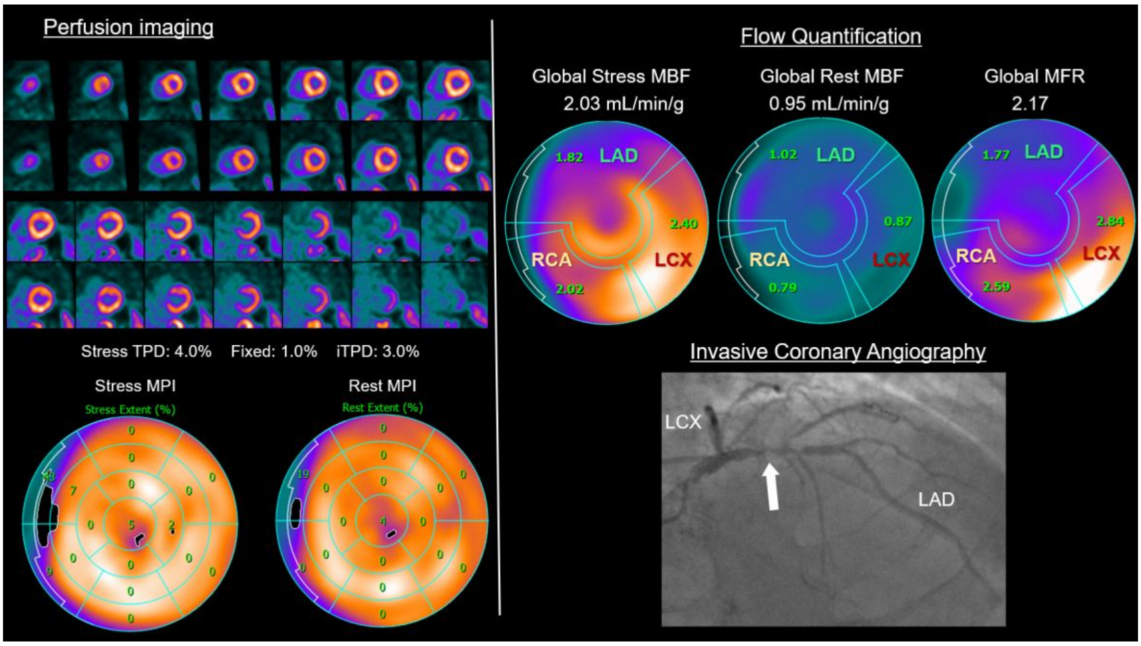

**Supplemental Figure 1. Clinical example of incremental value of regional MFR beyond global MFR**

A man in his 80s with hypertension and peripheral artery disease underwent Rubidium PET MPI for evaluation of dyspnea. Perfusion imaging showed an equivocal defect (ischemic total perfusion deficit [iTPD] 3%) in the LAD territory. Stress MBF and MFR in the LAD territory were reduced (1.82 mL/min/g and 1.77), whereas global stress MBF and MFR remained preserved (2.03 mL/min/g and 2.17). He was managed medically. Three years later, he developed left-sided chest and arm pain with mild troponin elevation. Coronary angiography revealed severe proximal LAD stenosis (white arrow) requiring primary PCI.

iTPD – ischemic total perfusion deficit

LAD – left anterior descending artery

LCX – left circumflex coronary artery

MBF – myocardial blood flow

MFR – myocardial flow reserve

MPI – myocardial perfusion imaging

PCI – percutaneous coronary intervention

PET – positron emission tomography

RCA – right coronary artery

TPD – total perfusion deficit

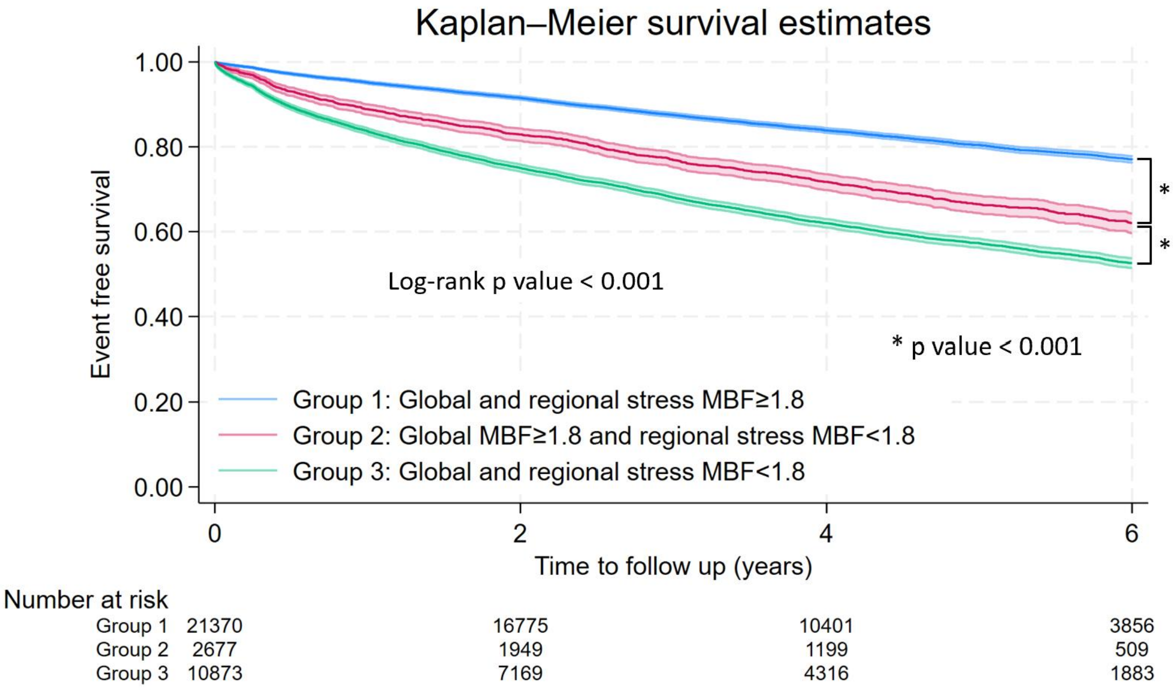

**Supplemental Figure 2.** Kaplan–Meier curves for MACE according to global and regional stress MBF

Kaplan–Meier estimates of major adverse cardiovascular events (MACE) according to global and regional myocardial blood flow (MBF). Event-free survival progressively declined across the 3 groups (log-rank $p$ < 0.001). Patients with preserved global but abnormal regional MBF had significantly worse outcomes than those with normal global and regional MBF.

MACE – major adverse cardiovascular event
MBF – myocardial blood flow
